# Enhanced Recovery After Cesarean from the Patient Perspective: A Prospective Study of the ERAC Questionnaire (ERAC-Q)

**DOI:** 10.1101/2023.06.07.23291068

**Authors:** Jessica A Meyer, Suzanne Alton, Hyunuk Seung, Autusa Pahlavan, Ariel R Trilling, Martha Coghlan, Katherine R Goetzinger, Liviu Cojocaru

## Abstract

**OBJECTIVE:** To evaluate the impact of an Enhanced Recovery After Cesarean (ERAC) protocol on the post-cesarean recovery experience using a validated ten-item questionnaire (ERAC-Q).

**METHODS:** This is a prospective cohort study of patients completing ERAC quality-of-life questionnaires (ERAC-Q) during inpatient recovery after cesarean delivery (CD) between October 2019 and September 2020, before and after the implementation of our ERAC protocol. Patients with non-Pfannenstiel incision, ICU admission, massive transfusion, bowel injury, existing chronic pain disorders, acute postpartum depression, or neonatal demise were excluded. The ERAC-Q was administered on postoperative day one and day of discharge to the pre– and post-ERAC implementation cohorts, rating aspects of their recovery experience on a scale of 0 (best) to 10 (worst). The primary outcome was ERAC-Q scores. Statistical analysis was performed with SAS software.

**RESULTS:** There were 196 and 112 patients in the pre– and post-ERAC cohorts, respectively. The post-ERAC group reported significantly lower total ERAC-Q scores compared to the pre-ERAC group, reflecting fewer adverse symptoms and greater perceived recovery on postoperative day one (1.6 [0.7, 2.8] vs. 2.7 [1.6, 4.3]) and day of discharge (0.8 [0.3, 1.5] vs. 1.4 [0.7, 2.2]) (p<0.001). ERAC-Q responses did not predict the time to achieve objective postoperative milestones. However, worse ERAC-Q pain and total scores were associated with higher inpatient opiate use.

**CONCLUSIONS:** ERAC implementation positively impacts patient recovery experience. The administration of ERAC-Q can provide real-time feedback on patient-perceived recovery quality and how healthcare protocol changes may impact their experience.

## INTRODUCTION

Standardized, multidisciplinary perioperative care through Enhanced Recovery After Surgery (ERAS) protocols have been consistently shown to reduce complication rates, minimize opioid use, and decrease the length of stay [1, 2]. There is now also a growing body of literature to show the qualitative benefits of these protocols on patient satisfaction and experience across a wide range of surgical specialties and procedure types [3–7]. Patient surveys, such as the Quality of Recovery (QoR) questionnaires, yield insights into some of the subjective outcomes that are of most value to patients, from perceived comfortability to psychological support [7, 8]. Examination of qualitative metrics via validated questionnaires allows for a more comprehensive understanding of the postoperative recovery experience from the patient’s perspective, informing the development of better perioperative care protocols [9–11].

In obstetrics, Enhanced Recovery After Cesarean (ERAC) protocols have been adapted from ERAS protocols to address some of the unique clinical challenges of postpartum recovery and pregnancy-specific complications [2, 12–17]. Compared to non-obstetric surgical patients, a post-cesarean patient is subject to unprecedented fluid shifts, hormonal fluctuations, marked hypercoagulability, and often pregnancy-specific therapies, such as magnesium for eclampsia prophylaxis [18]. To help address such unique issues, dedicated guidelines have been published by the ERAS Society to inform the antenatal, intraoperative, and postoperative care of cesarean delivery (CD) patients [19–21]. These guidelines are designed to target the same clinical objective outcomes that first inspired the original ERAS protocols but do not necessarily reflect the patient-perceived experiences of these interventions. Questionnaire evaluations of overall patient experiences after cesarean have been investigated, but the patient-perceived impact of enhanced recovery protocols for post-cesarean recovery is not well studied [8]. The aim of this study is to evaluate the impact of ERAC protocol implementation on the holistic post-cesarean patient recovery experience using a validated ten-item questionnaire (ERAC-Q). We hypothesized that the implementation of a mature ERAC protocol would improve the patient recovery experience.

## MATERIALS AND METHODS

This prospective cohort study was conducted at a single academic center to examine the impact of an ERAC protocol on the patient recovery experience among patients undergoing CD. The University of Maryland Institutional Review Board approved this study (#HP-00088872), and our ERAC protocol interventions were designed based on ERAS Society guidelines [19–21].

Pre-ERAC enrollment spanned from October 2019 to February 2020, while post-ERAC enrollment occurred from May 2020 to September 2020. The delayed post-ERAC enrollment period allowed for a two-month interval implementation phase of the ERAC protocol after the initial roll-out on the Obstetrical Care Unit (OBCU). Patients 18 years of age or older with a singleton gestation undergoing CD were considered for enrollment. Exclusion criteria were chosen to minimize factors that might significantly prolong recovery and/or increase postoperative opiate use [22]. Study population data were collected from electronic medical records and entered into a secure Research Electronic Data Capture (REDCap) database. This article was prepared in compliance with the Reporting on ERAS Compliance, Outcomes, and Elements Research (RECOvER) and the Strengthening the Reporting of Observational Studies in Epidemiology (STROBE) guidelines [23, 24].

ERAC interventions focused on four major areas: expansion of multimodal analgesia, reduction in perioperative fasting times, a standardized approach to early ambulation, and provision of patient educational visual aids on ERAC–based recovery (http://umm.edu/ERAC). Full details of our ERAC protocol and compliance with it are described in our prior published work [22].

The patient recovery experience was assessed via a ten-item questionnaire administered on postoperative day one and the day of discharge. It includes four questions evaluating postpartum symptom severity (pain, nausea/vomiting, dizziness, shivering) and six questions appraising patient recovery perceptions (overall comfortability and sense of control, independent mobility, activities of daily living, maternal-neonatal dyad interactions). For symptom-based questions, a zero score represents the lowest severity; for recovery-based questions, a zero score indicates the greatest degree of perceived recovery. For the development of the survey, we referenced and adapted previously validated questionnaires [25, 26]. Ease of reading was maintained using Flesch Kincaid grading. Visual analog was strengthened by the addition of emojis, which have a high level of agreement with the numeric rating scale [27]. The content and construct validity of the ERAC-Q were evaluated by the ERAC team [22, 28]. The questionnaire was made available in two languages (English and Spanish), with Spanish translation performed using professional services to ensure equivalence to the English version. The English version of our ERAC-Q is available in Appendix 1.

The primary outcome was ERAC-Q scores. Secondary outcomes included patient-reported mobility while on magnesium for eclampsia prophylaxis, correlation of ERAC-Q scores with the achievement of postoperative milestones (time to first feeding, time to ambulation, time to urinary catheter removal, time to hospital discharge), and correlation of ERAC-Q scores with opiate use in morphine milligram equivalents (MME).

Statistical analysis was performed with SAS (Version 9.4, SAS Institute, Cary, NC). Descriptive statistics were used for frequency, median, and mean, while associations between clinical characteristics and the ERAC implementation period (pre or post) were compared using Chi-square, Fisher’s exact, Wilcoxon rank-sum, or T-test where appropriate. Wilcoxon rank-sum test was used to assess the distribution of hospital MME use and ERAC questionnaire scores over the pre-ERAC and post-ERAC periods. The Mann-Whitney U test was used to examine relationships between categorical clinical outcomes and continuous ERAC questionnaire scores. Multiple logistic regression modeling was used to examine associations between inpatient MME use, ERAC questionnaire scores, and clinical factors. The selection of predictor variables was guided by descriptive statistics for categorical variables, Spearman correlation coefficients for continuous variables, and clinical relevance; variance inflation factors were calculated to facilitate the elimination of any multicollinearity. Goodness-of-fit and discriminatory power for this model were confirmed via the Hosmer-Lemeshow test and the area under the receiver operating characteristic curve (ROC). A similar assessment for the outcome of time to achieve postoperative milestones was conducted with a generalized linear model with gamma distribution. Statistical significance was defined at p<0.05.

## RESULTS

A total of 308 patients were enrolled in our study, including 196 in the pre-ERAC and 112 in the post-ERAC implementation arms. Patients in the post-ERAC cohort had a higher median BMI (36 vs. 32 pre-ERAC, p<0.001) and were more likely to have gestational diabetes (12% vs. 4% pre-ERAC, p<0.01), though less likely to have pregestational diabetes (4% vs. 13% pre-ERAC, p<0.01); baseline demographic and medical characteristics were otherwise similar between groups (Table 1). Surgical characteristics were also comparable (Supplemental Table 1). Postoperatively, the post-ERAC group achieved milestones, such as oral intake, urinary catheter removal, and ambulation, in less time than the pre-ERAC group (p<0.0001). There were no significant differences between groups in the frequency of surgical and delivery complications, intraoperative duration of CD, or estimated blood loss. There were also no significant differences in neonatal outcomes between groups (Supplemental Table 2).

**Table 1:** Demographic and medical characteristics of pre– and post-ERAC implementation cohorts

| Characteristic | Pre-ERAC<br>(n=196) | Post-ERAC<br>(n= 112) | p-value |
| --- | --- | --- | --- |
| Age, years | 30.6 ±6.4 | 30.7 ±5.5 | 0.9 |
| Race |  |  | 0.2 |
| Hispanic | 7 (3.6) | 10 (8.9) |  |
| Non-Hispanic Black | 121 (61.7) | 74 (66.1) |  |
| Non-Hispanic White | 56 (28.6) | 25 (22.3) |  |
| Asian | 8 (4.1) | 2 (1.8) |  |
| BMI, kg/m<sup>2</sup> | 32.1 [27.5, 39.5] | 35.8 [30.9, 41] | <0.001 |
| Parity | 1 [0, 2] | 1 [0, 1.5] | 0.1 |
| No pre-existing medical condition | 56 (28.6) | 16 (14.3) | 0.004 |
| Pregestational diabetes | 25 (12.8) | 4 (3.6) | 0.008 |
| Gestational diabetes | 7 (3.6) | 13 (11.6) | 0.006 |
| Chronic hypertension | 36 (18.4) | 25 (22.3) | 0.4 |
| Pregnancy-induced hypertension | 45 (23.0) | 16 (14.3) | 0.07 |
| Depression | 48 (24.5) | 21 (18.9) | 0.3 |
| Antidepressant use | 17 (35.4) | 5 (23.8) | 0.3 |
| Positive admission toxicology | 18 (9.2) | 13 (11.6) | 0.5 |
| Abbreviations: Enhanced Recovery After Cesarean (ERAC), Body Mass Index (BMI)<br>Data presented as n (%), mean ±SD, or median [IQR] where appropriate. |  |  |  |

ERAC-Q results varied between ERAC implementation cohorts. The overall survey response rate across two administration days per patient and both ERAC phases of the study was 64% (mean completion rate 71% post-ERAC vs. 59% pre-ERAC). Scores for nausea/vomiting, dizziness, and shivering did not vary significantly between ERAC cohorts, as the majority of patients in both groups did not report having these symptoms. However, self-reported pain symptoms and all recovery metrics were significantly improved for patients in the post-ERAC group, both on postoperative day one and the day of discharge (Table 2). Yet, among patients receiving magnesium therapy, perception of independent mobility (ERAC-Q #6) on postoperative day one was no longer significantly different between groups (score 1.5 [0, 5] post-ERAC vs. score 5 [4, 7] pre-ERAC, p=0.1). Throughout the hospital stay, the post-ERAC group reported significantly lower total ERAC-Q scores, reflecting fewer adverse symptoms and greater perceived recovery (postoperative day one total score 1.6 [0.7, 2.8] vs. 2.7 [1.6, 4.3] pre-ERAC; day of discharge total score 0.8 [0.3, 1.5] vs. 1.4 [0.7, 2.2] pre-ERAC).

**Table 2:** Distribution of ERAC-Q scores at the start and end of the inpatient postoperative recovery period in ERAC implementation cohorts

| Question<sup>a</sup> | Postoperative Day 1 |  |  | Day of Discharge |  |  |
| --- | --- | --- | --- | --- | --- | --- |
|  | Pre-ERAC<br>(n=116) | Post-ERAC<br>(n=86) | p-value | Pre-ERAC<br>(n=116) | Post-ERAC<br>(n=73) | p-value |
| **Q1** | 5 [4, 7.5] | 4 [2, 6] | 0.001 | 4 [3, 6] | 3 [2, 5] | 0.04 |
| **Q2** | 0 [0, 0] | 0 [0, 1] | 0.6 | 0 [0, 0] | 0 [0, 0] | 0.06 |
| **Q3** | 0 [0, 1] | 0 [0, 0] | 0.6 | 0 [0, 0] | 0 [0, 0] | 0.2 |
| **Q4** | 0 [0, 3] | 0 [0, 1] | 0.2 | 0 [0, 0] | 0 [0, 0] | 0.2 |
| **Q5** | 4 [2, 6] | 2 [0, 5] | 0.0001 | 3 [1, 5] | 1 [0, 4] | 0.007 |
| **Q6** | 5 [2, 7] | 1 [0, 5] | <0.0001 | 1 [0, 4] | 0 [0, 2] | <0.001 |
| **Q7** | 1 [0, 6] | 0 [0, 1] | 0.006 | 0 [0, 1] | 0 [0, 0] | 0.004 |
| **Q8** | 1 [0, 5] | 0 [0, 1] | 0.03 | 0 [0, 1] | 0 [0, 0] | 0.006 |
| **Q9** | 3 [0, 6] | 1 [0, 3] | 0.0001 | 1 [0, 2] | 0 [0, 1] | <0.001 |
| **Q10** | 3 [1, 6] | 0 [0, 2] | <0.0001 | 1 [0, 2] | 0 [0, 1] | <0.001 |
| **Total** | 2.7<br>[1.6, 4.3] | 1.6<br>[0.7, 2.8] | <0.0001 | 1.4<br>[0.7, 2.2] | 0.8<br>[0.3, 1.5] | <0.001 |

Abbreviations: Enhanced Recovery After Cesarean (ERAC)
Data presented as median [IQR].
<sup>a</sup>Q1 (severity of pain), Q2 (severity of nausea or vomiting), Q3 (severity of dizziness), Q4 (severity of shivering), Q5 (“I have been comfortable”), Q6 (“I can move independently”), Q7 (“I can hold my baby with no assistance”), Q8 (“I can feed/nurse my baby without assistance”), Q9 (“I can look after myself (shower, use the bathroom)”), Q10 (“I feel in control”).

ERAC-Q responses regarding postoperative pain improved significantly following ERAC protocol implementation (Table 2) and correlated with differences in MME use. Of all surveyed patients, median pain scores (ERAC-Q #1) and median total scores were higher among patients with inpatient MME use; no other ERAC-Q scores were found to be associated with significant variation in inpatient MME use or MME prescribed at discharge (Table 3). The significant predictive connection between pain score on the ERAC-Q and inpatient MME use persisted in multivariate analysis, even after adjusting for maternal depression, positive admission toxicology, and delivery or surgical complications (OR 1.44, 95% CI 1.2-1.73, p<0.0001) (Table 4). Multivariate analyses did not demonstrate a similar predictive association between ERAC-Q scores and time to achieving objective postoperative milestones (ambulation, oral intake, and discharge); in adjusting for other clinical factors, logistic regression modeling did reveal a 13% increase in average time to discharge with the presence of positive maternal toxicology on admission (p<0.01) and a 38% increase in the average time to ambulation in the setting of any delivery complication (p=0.03) (Table 5).

**Table 3:** Distribution of discharge day ERAC-Q scores and associated MME

| Question<sup>a</sup> | Any Inpatient MME Use |  |  | Any Prescribed Outpatient MME |  |  |
| --- | --- | --- | --- | --- | --- | --- |
|  | No<br>(n=41) | Yes<br>(n=148) | p-value | No<br>(n=16) | Yes<br>(173) | p-value |
| **Q1** | 2 [1, 4] | 4 [3, 7] | <0.0001 | 3.5 [2, 6.5] | 4 [2, 6] | 0.5 |
| **Q2** | 0 [0, 0] | 0 [0, 0] | 0.2 | 0 [0, 0.5] | 0 [0, 0] | 0.2 |
| **Q3** | 0 [0, 0] | 0 [0, 0] | 0.3 | 0 [0, 0] | 0 [0, 0] | 0.9 |
| **Q4** | 0 [0, 0] | 0 [0, 0] | 0.7 | 0 [0, 0] | 0 [0, 0] | 0.5 |
| **Q5** | 1 [0, 4] | 2 [0, 5] | 0.3 | 2.5 [0, 4] | 2 [0, 5] | 0.7 |
| **Q6** | 1 [0, 2] | 1 [0, 3] | 0.3 | 0.5 [0, 2] | 1 [0, 3] | 0.4 |
| **Q7** | 0 [0, 1] | 0 [0, 1] | 0.9 | 0 [0, 1] | 0 [0, 1] | 1.0 |
| **Q8** | 0 [0, 1] | 0 [0, 1] | 0.5 | 0.5 [0, 1] | 0 [0, 1] | 0.6 |
| **Q9** | 0 [0, 1] | 0 [0, 2] | 0.4 | 0 [0, 1] | 0 [0, 1] | 1.0 |
| **Q10** | 0 [0, 1] | 0 [0, 2] | 0.9 | 1 [0, 3.5] | 0 [0, 2] | 0.1 |
| **Total Score** | 0.8<br>[0.3, 1.3] | 1.3<br>[0.6, 2] | 0.009 | 1.2<br>[0.5, 1.9] | 1.3<br>[0.5, 2] | 0.8 |

Abbreviations: Enhanced Recovery After Cesarean (ERAC), Morphine Milligram Equivalents (MME)
Data presented as median [IQR].
<sup>a</sup>Q1 (severity of pain), Q2 (severity of nausea or vomiting), Q3 (severity of dizziness), Q4 (severity of shivering), Q5 ("I have been comfortable"), Q6 ("I can move independently"), Q7 ("I can hold my baby with no assistance"), Q8 ("I can feed/nurse my baby without assistance"), Q9 ("I can look after myself (shower, use the bathroom)"), Q10 ("I feel in control").

**Table 4:** Multivariate analysis for inpatient MME use in the overall study population

| Predictor Variable | Adjusted OR (95% CI) | p-value |
| --- | --- | --- |
| ERAC Q1 Score<sup>a</sup> | 1.44(1.2, 1.73) | <0.001 |
| Depression<sup>b</sup> | 0.94(0.34, 2.56) | 0.9 |
| Positive admission toxicology<sup>b</sup> | 2.5(0.51, 12.23) | 0.3 |
| Any delivery complication<sup>b</sup> | 7.82(0.95, 64.23) | 0.06 |
| Any surgical complication<sup>b</sup> | 0.95(0.41, 2.19) | 0.9 |
| Abbreviations: Morphine Milligram Equivalents (MME), Enhanced Recovery After Cesarean (ERAC), Odds Ratio (OR)<br><sup>a</sup>ERAC Q1 scores for severity of pain on postoperative day one and/or day of discharge.<br><sup>b</sup>Regression model predictor references are absence of depression history, negative admission toxicology, absence of delivery complication, and absence of surgical complication, respectively. |  |  |

**Table 5:** Multivariate analysis for time to achieve postoperative milestones in the overall study population

|  | Predictor Variable | Estimate (SE) | % Mean Difference (95% CI) | p-value |
| --- | --- | --- | --- | --- |
| Time to ambulation | ERAC Q3 Score<sup>a</sup> | 0.06 (0.04) | 5.8 (-1.3, 13.3) | 0.1 |
|  | ERAC Q8 Score<sup>a</sup> | 0.05 (0.03) | 5.1 (-1.3, 11.8) | 0.1 |
|  | Depression<sup>a</sup> | -0.13 (0.12) | -12.2 (-30.8, 11.7) | 0.3 |
|  | Positive admission toxicology<sup>b</sup> | 0.08 (0.16) | 8.1 (-21.4, 48.7) | 0.6 |
|  | Any delivery complication<sup>b</sup> | 0.32 (0.15) | 37.7 (2.9, 84.1) | 0.03 |
|  | Any surgical complication<sup>b</sup> | -0.16 (0.10) | -14.8 (-30.4, 3.5) | 0.1 |
| Time to feeding | ERAC Q2 Score<sup>a</sup> | 0.12 (0.11) | 12.7 (-8.6, 38.9) | 0.3 |
|  | ERAC Q3 Score<sup>a</sup> | 0.11 (0.07) | 11.6 (-2.6, 29.1) | 0.1 |
|  | ERAC Q8 Score<sup>a</sup> | 0.07 (0.07) | 7.4 (-7, 23.9) | 0.3 |
|  | ERAC Q10 Score<sup>a</sup> | 0.01 (0.06) | 1 (-9.4, 12.5) | 0.9 |
|  | Depression<sup>a</sup> | -0.17 (0.24) | -15.6 (-46.9, 33.9) | 0.5 |
|  | Positive admission toxicology<sup>b</sup> | 0.14 (0.29) | 15 (-34.5, 102.3) | 0.6 |
|  | Any delivery complication<sup>b</sup> | 0.1 (0.28) | 10.5 (-35.6, 91.2) | 0.7 |
|  | Any surgical complication<sup>b</sup> | -0.24 (0.18) | -21.3 (-45.4, 12.4) | 0.2 |
| Time to discharge | ERAC Q1 Score<sup>a</sup> | 0.006 (0.006) | 0.6 (-0.5, 1.7) | 0.3 |
|  | ERAC Q10 Score<sup>a</sup> | 0.01 (0.006) | 1.1 (-0.1, 2.3) | 0.08 |
|  | Depression<sup>a</sup> | -0.005 (0.03) | -0.5 (-6.9, 6.5) | 0.9 |
|  | Positive admission toxicology<sup>b</sup> | 0.12 (0.05) | 12.7 (3.5, 23.6) | 0.007 |
|  | Any delivery complication<sup>b</sup> | 0.04 (0.04) | 4.2 (-3.8, 12.9) | 0.3 |
|  | Any surgical complication<sup>b</sup> | 0.04 (0.03) | 4.2 (-1.6, 10.3) | 0.2 |

Abbreviations: Enhanced Recovery After Cesarean (ERAC)
<sup>a</sup>ERAC-Q scores collected on postoperative day one and/or day of discharge. Q1 (severity of pain), Q2 (severity of nausea or vomiting), Q3 (severity of dizziness), Q8 (“I can feed/nurse my baby without assistance”), Q10 (“I feel in control”).
<sup>b</sup>Regression model predictor references are absence of depression history, negative admission toxicology, absence of delivery complication, and absence of surgical complication, respectively.

ERAC implementation was associated with more optimal recovery scores on the ERAC-Q by day of discharge. Responses to most questions differed significantly between the pre– and post-ERAC cohorts in the frequency of a score zero. A zero score (greatest degree of perceived recovery) on day of discharge was more common in the post-ERAC group for symptoms of nausea/vomiting and for all included recovery metrics (Table 6). Discharge day questionnaire responses in each ERAC cohort did not significantly differ by neonatal NICU admission or by subsequent maternal hospital readmission and postpartum visit attendance (Supplemental Table 3).

**Table 6:** Distribution of ERAC questionnaire questions with score zero on day of discharge in ERAC implementation cohorts

| **Question<sup>a</sup>**<br>**with Score Zero** | **Pre-ERAC**<br>**(n=116)** | **Post-ERAC**<br>**(n=73)** | **p-value** |
| --- | --- | --- | --- |
| **Q1** | 9 (7.9)<sup>b</sup> | 8 (11.0) | 0.5 |
| **Q2** | 96 (82.8) | 68 (93.2) | 0.04 |
| **Q3** | 99 (85.3) | 67 (91.8) | 0.2 |
| **Q4** | 100 (86.2) | 67 (91.8) | 0.2 |
| **Q5** | 21 (18.3)<sup>b</sup> | 33 (45.2) | <0.001 |
| **Q6** | 34 (29.6)<sup>b</sup> | 44 (60.3) | <0.001 |
| **Q7** | 38 (55.1)<sup>b</sup> | 36 (80)<sup>b</sup> | 0.006 |
| **Q8** | 37 (55.2)<sup>b</sup> | 36 (80)<sup>b</sup> | 0.007 |
| **Q9** | 54 (47.0)<sup>b</sup> | 54 (74.0) | <0.001 |
| **Q10** | 47 (40.5) | 54 (74.0) | <0.001 |
| **Total** | 6 (5.2) | 6 (8.2) | 0.5 |
| Abbreviations: Enhanced Recovery After Cesarean (ERAC)<br>Data presented as n (%).<br><sup>a</sup>Q1 (severity of pain), Q2 (severity of nausea or vomiting), Q3 (severity of dizziness), Q4 (severity of shivering), Q5 (“I have been comfortable”), Q6 (“I can move independently”), Q7 (“I can hold my baby with no assistance”), Q8 (“I can feed/nurse my baby without assistance”), Q9 (“I can look after myself (shower, use the bathroom)”), Q10 (“I feel in control”).<br><sup>b</sup>Q1: Pre-ERAC (n=114); Q5 and Q6 and Q9: Pre-ERAC (n=115); Q7: Pre-ERAC (n=69); Q7 and Q8: Post-ERAC (n=45); Q8: Pre-ERAC (n=67) |  |  |  |

## DISCUSSION

In this prospective cohort study, we implemented an ERAC protocol at a single academic center’s OBCU and examined its impact on subjective aspects of the patient recovery experience using a validated questionnaire. With heavy emphasis on patient education, our protocol targeted preoperative preparation, expanded postoperative non-opiate analgesia, and standardized the approach to postoperative goal setting. Responses to a ten-item questionnaire (ERAC-Q) revealed significant patient-reported improvements after ERAC implementation, including lower pain scores and greater perceived recovery across all surveyed metrics. This demonstrates that a standardized, multidimensional care protocol can effect positive change on the patient-reported overall recovery experience.

A CD is a major abdominal surgery, but compounding the challenges of postoperative healing is the fact that the birthing process alone is a massive adjustment, regardless of delivery mode. As some postpartum physiology poses maternal health risks, continued optimization of ERAC protocols should take care to include standardized interventions that address them. Further, delivery complications and peripartum treatments that are unique to obstetrics, such as magnesium for seizure prophylaxis in preeclamptic patients, can also influence outcomes and patient perceptions of recovery. In our prior work, we found that the objective time to ambulation postpartum was more than twice as long for patients receiving eclampsia prophylaxis with magnesium, and the beneficial effect of ERAC on time to achieve this milestone was dulled [22]. In the current study, we found that patients’ perceptions of mobility likewise were negatively impacted by magnesium therapy, despite the benefits of ERAC protocol implementation shown in the cohorts not treated with magnesium. Complex obstetrical circumstances would benefit from the building of more comprehensive ERAC algorithms to standardize care plans for scenarios in which deviation from the routine ERAC protocol may be necessary or clinically unavoidable.

Even more pervasive than the physical changes are the social, mental, and emotional adjustments that accompany birth and dramatically influence the patient recovery experience [10, 11, 29]. A substantial number of women in a US survey reported new health concerns postpartum related to adjusting to motherhood [11, 30]. Investment in the needs of a neonate, the struggle to prioritize self-care, and unforeseen relationship changes are only a few of the fourth-trimester adjustments for which patients often report feeling unprepared [11, 29, 30]. Further, unpredictable birth events, such as emergent, unplanned CD, also likely contribute to the themes of dissatisfaction, failure, lack of control, and poor neonatal bonding reported in several qualitative post-cesarean studies [9, 30, 31]. This underscores the value of patient-reported outcome and experience measures (PROMs and PREMs), such as those included in our ERAC-Q. As there is often a discrepancy between the immediate recovery concerns of clinicians and their postpartum patients, active attention to the patient voice, particularly among marginalized groups, is critical to identifying opportunities for improving care [11, 30, 32]. Post-cesarean patients in an open interview-based study of their ERAC experiences reported that earlier phase-specific education, improved provider communication in the perioperative period, and greater emphasis on skin-to-skin bonding would have greatly improved their recovery care [33]. Modification of ERAC protocols to incorporate decision aides throughout the birthing process and structured opportunities for shared decision-making may help to better marry the different aims of providers and patients for a more holistic quality care [32].

Our study found a significant improvement in patients’ perceptions of their recovery after ERAC implementation across multiple elements of care, including pain control, comfort, feelings of control, and independent function for neonatal and self-care. This is consistent with the findings of a smaller ERAC questionnaire study that administered the QoR15 questionnaire on the day of discharge [34]. Our study implemented obstetric-specific patient recovery perceptions and was administered at the beginning and end of the postpartum recovery period to assess patient experiences over time. In our study, we suspect that the significant improvements in patient-reported metrics might be due in large part to our ERAC protocol’s emphasis on patient education (http://umm.edu/ERAC). Both antenatally and perioperatively, our protocol prioritized instructing patients on preparation for CD, setting expectations for intraoperative events and the ERAC protocol, as well as describing possible complications for surveillance at home. The benefits of patient education initiatives for patient satisfaction and experience are well-studied in the literature [31, 35–38]. Our study further supports the assertion that an ERAC protocol with a dedicated educational component can help to restore patients’ sense of control and well-being following a cesarean birth.

Our prospective study exhibits several strengths, most notably its attention to the patient-reported experience of recovery before and after ERAC protocol implementation, which is currently not studied. While objective postoperative milestones are undeniably important to recovery, our survey assessed patient perceptions of a wider range of valuable post-cesarean goals, adding depth of understanding to the patient experience. As the circumstances behind delivery can greatly impact a patient’s experience, an added strength of our study is the inclusion of non-elective CD, which allows for a wider range of delivery scenarios. Additionally, the patient diversity of our study population facilitates generalizability with excellent representation of Hispanic and non-Hispanic Black patients, which is particularly important as ERAC protocols have been found to help address racial disparities in healthcare [39]. Lastly, our survey is a patient quality-of-life-after-cesarean survey and was administered on repeat occasions during the inpatient admission to evaluate changes in patient-reported recovery semi-longitudinally. We used both numeric and visual analog scales in the ERAC-Q, as well as translation into the two most spoken languages in the United States, to aid in maximizing usability and response reliability [40–42].

There are also several limitations to our work. Considerable efforts were made to facilitate ERAC protocol implementation and encourage adherence in the OBCU. While helpful for ensuring the practical success of a new care model integration, this may also introduce performance bias that could impact observed differences between cohorts. This might be reflected in the significant improvement in the survey response rate for the post-ERAC group, though the overall response rate was similar to national rates [43]. Moreover, recent literature suggests that lower response rates may have less influence on outcomes than previously thought [44, 45]. Another limitation of our study relates to patient recovery after discharge. Data was collected on MME prescribed at discharge and rates of readmission or postpartum visit attendance, but more granular details about patients’ recovery experiences at home (for example – outpatient MME consumption, length of maternity leave, social support, or breastfeeding success) remain unexplored due to lack of longitudinal follow-up in our study. Further, our ERAC-Q is a quality-of-life survey, not a patient satisfaction survey, which would examine congruence with patients’ expectations of their care (such as interactions with providers or the conditions of the hospital environment). To critically assess protocol performance, patient satisfaction measures should also be examined as these reflect care adequacy, independent of patient-reported quality of life [3, 5, 6, 46–48]. Future research on ERAC should focus on examining more patient-reported metrics for both inpatient and outpatient recovery to continue improving care quality and adequacy.

The findings of our study reflect the impact of a standardized ERAC protocol on the multidimensional process of patient recovery after CD. Efforts to build upon and further integrate such protocols must ensure dedicated attention to the unique risks and demands of the postoperative and postpartum states.

Further, optimal post-cesarean care must also integrate the more subjective insights provided by patient-reported measures into developing patient-centered ERAC protocols. Routine administration of patient questionnaires, such as ERAC-Q, can provide real-time feedback regarding patient-perceived recovery quality and the impact that protocol changes may have on their experience.

## Data Availability

All data produced in the present study are available upon reasonable request to the authors.

---

Supplemental Table 1: Perioperative characteristics of pre– and post-ERAC implementation cohorts

(see separately uploaded supplementary materials)

Supplemental Table 2: Neonatal characteristics of ERAC implementation cohorts (see separately uploaded supplementary materials)

Supplemental Table 3: Distribution of discharge day ERAC questionnaire scores by postpartum visit attendance, inpatient readmission, and neonatal NICU admission (see separately uploaded supplementary materials)

Appendix 1: English Enhanced Recovery After Cesarean Questionnaire (ERAC-Q)

(see separately uploaded supplementary materials)

