## Supplemental Tables and Appendix for "Enhanced Recovery After Cesarean from the Patient Perspective: A Prospective Study of the ERAC Questionnaire (ERAC-Q)"

Supplemental Table 1: Perioperative characteristics of pre- and post-ERAC implementation cohorts

| Characteristic | Pre-ERAC (n=196) | Post-ERAC (n= 112) | p-value |
|---|---|---|---|
| Prior cesarean delivery | 91(46.4) | 53(47.3) | 0.9 |
| Prior laparotomy | 8 (4.1) | 11 (9.8) | 0.04 |
| GA at CD, days | 268 [251, 276] | 268 [256, 274] | 0.7 |
| Indication for CD | | | |
| Elective | 59 (30.1) | 43 (38.4) | 0.1 |
| Labor arrest | 22 (11.2) | 18 (16.1) | 0.2 |
| NRFHT | 39 (19.9) | 25 (22.3) | 0.6 |
| Malpresentation | 24 (12.2) | 11 (9.8) | 0.5 |
| Maternal indication | 37 (18.9) | 16 (14.3) | 0.3 |
| Placental abnormality | 12 (6.1) | 1 (0.9) | 0.04 |
| Blood loss, mL | 763 [507, 1047] | 751 [522, 1123] | 0.9 |
| CD operative time, min | 64.5 [52, 88] | 64.5 [51, 82.5] | 0.6 |
| No delivery complication | 171 (87.2) | 91 (81.3) | 0.2 |
| PPH | 62 (31.6) | 35 (31.3) | 0.9 |
| Chorioamnionitis | 9 (4.6) | 5 (4.5) | 1.0 |
| Incision extension | 11 (5.6) | 2 (1.8) | 0.1 |
| No surgical complication | 133 (67.9) | 74 (66.1) | 0.7 |
| SSI | 3 (1.5) | 1 (0.9) | 1.0 |
| Time to ambulation, hrs | 16 [11, 22.6] | 9.5 [6.6, 16.9] | <0.001 |
| Time to urinary foley removal, hrs | 12.5 [10.9, 17.8] | 10.1 [7.2, 13.7] | <0.001 |
| Time to feeding, hrs | 9.7 [6.8, 12.9] | 2.1 [1, 6] | <0.001 |
| Time to discharge, hrs | 76.1 [69.9, 90] | 72.4 [62.7, 82.7] | 0.002 |

Abbreviations: Enhanced Recovery After Cesarean (ERAC), Gestational Age (GA), Cesarean Delivery (CD), Non-reassuring Fetal Heart Tracing (NRFHT), Postpartum Hemorrhage (PPH, blood loss ≥1L), Surgical Site Infection (SSI)
Data presented as n (%), mean ±SD, or median [IQR] where appropriate.

Supplemental Table 2: Neonatal characteristics of ERAC implementation cohorts

| Characteristic | Pre-ERAC (n=196) | Post-ERAC (n= 112) | p-value |
|---|---|---|---|
| Neonatal sex | | | 0.9 |
| Male | 97 (49.5) | 54 (48.2) | |
| Female | 99 (50.5) | 58 (51.8) | |
| Birth weight, g | 3110 [2370, 3455] | 3163 [2667, 3525] | 0.1 |
| APGAR at 1 min | 8 [7, 8] | 8 [8, 8] | 0.8 |
| APGAR at 5 mins | 9 [9, 9] | 9 [9, 9] | 0.6 |
| pH umbilical artery | 7.22 [7.17, 7.27] | 7.22 [7.18, 7.26] | 0.4 |
| BE umbilical artery | -4.15 [-6.3, -2.75] | -4.2 [-6.5, -2.9] | 0.8 |
| pH umbilical vein | 7.29 [7.24, 7.33] | 7.29 [7.25, 7.32] | 0.6 |
| BE umbilical vein | -3.5 [-5.3, -2.1] | -3.2 [-4.7, -2.3] | 0.6 |
| NICU admission | 76 (38.8) | 37 (33.0) | 0.2 |

Abbreviations: Enhanced Recovery After Cesarean (ERAC), Base Excess (BE), Neonatal Intensive Care Unit (NICU)

Data presented as n (%) or median [IQR] where appropriate.

Supplemental Table 3: Distribution of discharge day ERAC questionnaire scores by postpartum visit attendance, inpatient readmission, and neonatal NICU admission

| | Postpartum Visit | | Readmission | | NICU | |
|---|---|---|---|---|---|---|
| | No (n=28) | Yes (n=161) | No (n=179) | Yes (n=10) | No (n=115) | Yes (n=72) |
| Q1[a] | 4 [2, 6.5] | 4 [2, 6] | 4 [2, 6] | 5 [4, 7] | 4 [2, 6] | 4 [2.5, 6] |
| Q2[a] | 0 [0, 0] | 0 [0, 0] | 0 [0, 0] | 0 [0, 0] | 0 [0, 0] | 0 [0, 0] |
| Q3[a] | 0 [0, 0] | 0 [0, 0] | 0 [0, 0] | 0 [0, 0] | 0 [0, 0] | 0 [0, 0] |
| Q4[a] | 0 [0, 0] | 0 [0, 0] | 0 [0, 0] | 0 [0, 0] | 0 [0, 0] | 0 [0, 0] |
| Q5[a] | 2 [0, 5] | 2 [0, 5] | 2 [0, 5] | 4 [1, 6] | 2 [0, 5] | 2 [0, 5] |
| Q6[a] | 1 [0, 2] | 1 [0, 3] | 1 [0, 3] | 0.5 [0, 3] | 1 [0, 3] | 1 [0, 3] |
| Q7[a] | 0 [0, 1] | 0 [0, 1] | 0 [0, 1] | 0 [0, 1] | 0 [0, 1] | na |
| Q8[a] | 0 [0, 0.5] | 0 [0, 1] | 0 [0, 1] | 1 [0, 3] | 0 [0, 1] | na |
| Q9[a] | 0 [0, 1] | 0 [0, 1] | 0 [0, 1] | 0 [0, 2] | 0 [0, 1] | 0 [0, 2] |
| Q10[a] | 1 [0, 3] | 0 [0, 1] | 0 [0, 2] | 0 [0, 4] | 0 [0, 1] | 0 [0, 3] |
| Total Score | 1.2 [0.5, 1.8] | 1.3 [0.5, 2] | 1.2 [0.5, 1.9] | 1.4 [1.1, 2.2] | 1.3 [0.6, 2] | 1.3 [0.5, 2.3] |

Abbreviations: Enhanced Recovery After Cesarean (ERAC), Neonatal Intensive Care Unit (NICU)

Data presented as median [IQR].

[a]Q1 (severity of pain), Q2 (severity of nausea or vomiting), Q3 (severity of dizziness), Q4 (severity of shivering), Q5 (“I have been comfortable”), Q6 (“I can move independently”), Q7 (“I can hold my baby with no assistance”), Q8 (“I can feed/nurse my baby without assistance”), Q9 (“I can look after myself (shower, use the bathroom)”), Q10 (“I feel in control”).

### Appendix 1: English Enhanced Recovery After Cesarean Questionnaire (ERAC-Q)

| ERAC Q score | Day 1 | Day of discharge | Label |
|---|---|---|---|

Below is a list of symptoms that women can experience after childbirth.
Please read each of them carefully and give a score from 0 to 10 for each symptom.

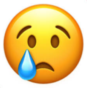
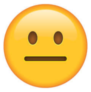
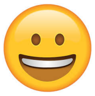

| **Symptoms score** | | Worst imaginable | | | | | Moderate | | | | | None |
|---|---|---|---|---|---|---|---|---|---|---|---|---|
| | | **10** | **9** | **8** | **7** | **6** | **5** | **4** | **3** | **2** | **1** | **0** |
| 1 | **Pain** | | | | | | | | | | | |
| 2 | **Nausea or vomiting** | | | | | | | | | | | |
| 3 | **Dizziness** | | | | | | | | | | | |
| 4 | **Shivering** | | | | | | | | | | | |

Please score your recovery in the last 24 hours. Please give a score from 0 to 10 for each aspect separately.

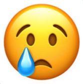
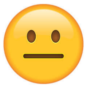
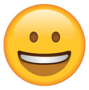

| **Recovery score** | | No/ Never | | | | | Sometimes or with help | | | | | Yes/ always |
|---|---|---|---|---|---|---|---|---|---|---|---|---|
| | | **10** | **9** | **8** | **7** | **6** | **5** | **4** | **3** | **2** | **1** | **0** |
| 5 | **I have been comfortable** | | | | | | | | | | | |
| 6 | **I can move independently** | | | | | | | | | | | |
| 7 | **I can hold my baby without assistance** | | | | | | | | | | | |
| 8 | **I can feed/ nurse my baby without assistance** | | | | | | | | | | | |
| 9 | **I can look after myself (shower, use the bathroom)** | | | | | | | | | | | |
| 10 | **I feel in control** | | | | | | | | | | | |

Comments:
